# Nutritional education program counseling based on the Mediterranean diet on intestinal constipation in people with Parkinson’s disease: a randomized clinical trial

**DOI:** 10.64898/2026.05.07.26352682

**Authors:** Raissa Dias Fernandez, Franciele da Silva Mesquita, Gabriella Cunha Ferreira, Bruno Lopes Santos-Lobato

## Abstract

**Introduction:** Intestinal constipation (IC) is a common and early non-motor symptom in Parkinson’s disease (PD), impacting patients’ quality of life. In this context, the Mediterranean diet plays a fundamental role in managing IC. This study aimed to evaluate the effects of a nutritional education program based on the principles of the Mediterranean diet on IC in individuals with PD.

**Methods:** This is a randomized, controlled, single-center, parallel-group, single-blind clinical study to evaluate the effectiveness of a nutritional education program based on the Mediterranean diet for three months, with a delayed-start design, in people with PD and IC. Participants were randomly allocated (1:1 ratio) to two groups: early-start (intervention from baseline to the third month) and delayed-start (intervention from the third to the sixth month) nutritional counseling, with an initial in-person assessment and monthly remote assessments. Bowel habits, adherence to the Mediterranean diet, and clinical variables were analyzed.

**Results:** After six months, a significant increase in the frequency of weekly bowel movements was observed (Early-start: 2.91 to 4.14; Delayed-start: 2.68 to 4.18 bowel movements/week; p < 0.001), along with changes in stool consistency and improved adherence to the Mediterranean diet over time. However, no significant differences were detected between the groups.

**Conclusion:** A nutritional education program based on the principles of the Mediterranean diet was associated with improved bowel habits and dietary adherence over time. These results support that locally adapted, low-cost dietary counseling may represent a complementary approach to the treatment of IC in individuals with PD in non-Mediterranean settings.

## 1. Introduction

Parkinson’s disease (PD) is a progressive neurodegenerative disorder characterized by the accumulation of pathological α-synuclein, a protein implicated in the formation of Lewy bodies and neurites, which are involved in both genetic and sporadic forms of the disease [1,2,3]. PD is characterized by motor symptoms (resting tremor, muscular rigidity, bradykinesia, and postural instability) and non-motor symptoms, including depression, anxiety, sleep disorders, autonomic dysfunction, and olfactory impairment [3]. Among non-motor symptoms, intestinal constipation (IC) is among the most prevalent comorbidities in PD, which may affect up to 80% of affected people [4,5,6].

There is bidirectional communication between the brain and the gastrointestinal tract mediated by the gut–brain axis, which involves interactions among the central, autonomic, and enteric nervous systems. Alterations in this axis may impair gastrointestinal motility, contributing to the development of IC in people with PD [5,6]. In addition, evidence indicates that PD is associated with peripheral and central nervous system inflammation, as well as increased intestinal permeability, reinforcing the relevance of the gastrointestinal tract in the pathophysiology of the disease [7].

In this context, diet plays a fundamental role in the management of IC. Adequate intake of dietary fiber, derived from fruits, vegetables, and whole grains, promotes intestinal motility, modulation of the gut microbiota, and the production of short-chain fatty acids, which exert anti-inflammatory effects and support intestinal mucosal integrity [8]. The Mediterranean diet is characterized by a dietary pattern rich in natural or minimally processed foods and fiber, and it is widely recognized for its anti-inflammatory properties and metabolic benefits [5,9]. However, this type of diet may be less affordable in certain regions due to cultural, geographic, and economic factors. Economic constraints play a fundamental role in food choices, often limiting access to foods commonly recommended in the Mediterranean diet, such as fruits, vegetables, and nuts [10,11]. In the Brazilian Amazon, traditional dietary habits include region-specific foods such as cassava flour and açaí, which represent important cultural and nutritional components of the local diet and are also widely available and low-cost [12].

Therefore, adapting the Mediterranean diet principles to regionally available, culturally relevant foods may be a feasible strategy to improve dietary adherence and promote better health outcomes. Based on this, the aim of this study was to assess the efficacy of a nutritional education program counseling based on Mediterranean diet principles on IC in people with PD.

## 2. Methods

### 2.1. Study design and participants

This was a parallel-group, monocentric, single-blind, randomized controlled trial to assess the efficacy of a Mediterranean diet-based nutritional education program delivered via counseling over three months in people with PD and IC. This study was submitted and approved by the Research Ethics Committee of Hospital Ophir Loyola, in accordance with Resolution 466/12 of the National Health Council, under approval number 6.611.167, prior to the initiation of participant contact and data collection. All participants received detailed information regarding the study objectives and procedures and signed an informed consent form. The trial was also registered in the Brazilian Registry of Clinical Trials (ReBEC) under registration number RBR-36q469s.

From January 2024 to December 2025, people with PD diagnosed according to the United Kingdom Parkinson’s Disease Society Brain Bank criteria [13] were screened from the Movement Disorders Clinic at Hospital Ophir Loyola (Belém, Brazil). Recruitment occurred during routine follow-up visits, and participation was voluntary and did not interfere with participants’ clinical treatment. The inclusion criteria were as follows: (i) age 18 years or older, (ii) diagnosis of IC according to stool types 1 or 2 on the Bristol Stool Form Scale validated for the Brazilian population [14] and with chronic IC according to the Rome III Criteria. According to the Rome III Criteria, chronic IC is defined as the presence of at least two criteria in ≥25% of bowel movements for a minimum of 12 weeks in recent months [15]. The exclusion criteria were as follows: (i) clinical diagnosis of dementia, (ii) presence of other neurological diseases, (iii) use of pharmacological laxatives (such as stimulant and osmotic laxatives), and (iv) diagnosis or history of previous inflammatory or neoplastic gastrointestinal diseases.

### 2.2. Randomization

Participants were randomly assigned (1:1 ratio) to two groups: early-start Mediterranean diet-based nutritional counseling (Early-start) and delayed-start nutritional counseling (Delayed-start). Randomization was performed using a web-based device, stratified by sex and disease duration, and was conducted by an independent researcher who did not participate in recruitment, assessment, or data analysis. The researchers responsible for in-person outcome assessments or data analysis remained blinded to group allocation. An independent group of researchers was responsible for delivering the nutritional counseling and had access to the group allocation but did not participate in outcome assessments. Due to the nature of the intervention, participants and the researchers responsible for delivering the nutritional education could not be blinded to the treatment allocation.

### 2.3. Procedures

The summary of assessments and interventions in the study is illustrated in Fig. 1. Participants were evaluated in person for clinical outcomes at baseline (T0), after three months (T3), and after six months (T6) from baseline by the same blinded researcher (principal investigator). At baseline, all participants were submitted to a socioeconomic questionnaire, and the clinical assessment of PD was performed using the Movement Disorder Society–Unified Parkinson’s Disease Rating Scale (MDS-UPDRS) [16] and the Hoehn and Yahr scale [17]. Phone calls were performed after one month (T1), two months (T2), four months (T4), and five months (T5) from baseline to provide support, ensure adherence during the intervention, and assess outcomes by a non-blinded researcher. Participants were not allowed to receive any other nutritional counseling during the study period.

**Fig. 1.**
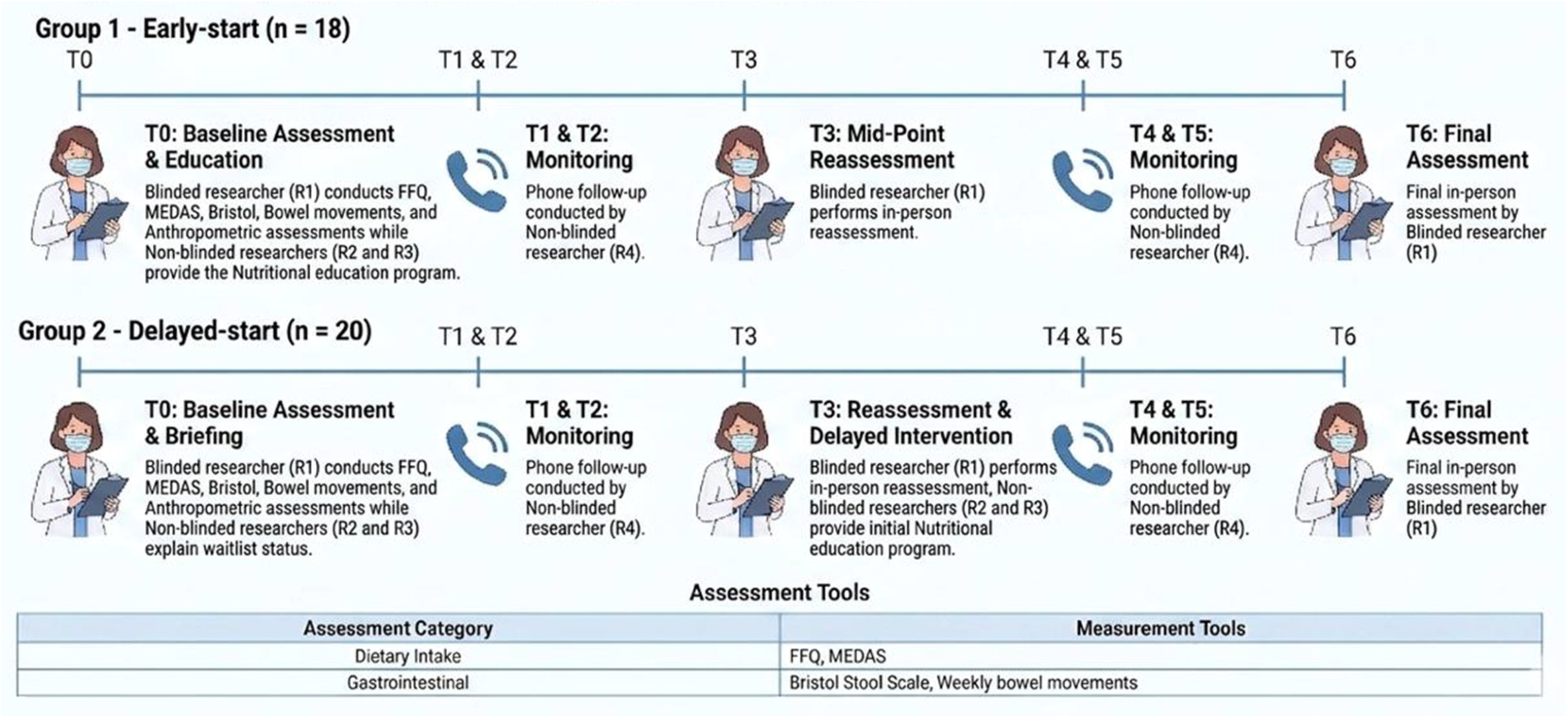
Study design and assessment protocol. In-person assessments were performed at T0, T3, and T6, with monthly telephone monitoring at T1, T2, T4, and T5.

In the nutritional education program counseling, participants were advised to include the following foods: nuts, eggs, fish, seafood, low-fat cheese, and whole grains. They were also instructed to limit the consumption of cream, butter/margarine, red and processed meats, carbonated and sugar-sweetened beverages, baked goods, and industrialized desserts, as well as French fries or potato chips. The education program also included recommendations on hydration and modifications to defecatory posture, such as using a footstool during toilet use.

The educational program was printed and delivered to participants (Supplementary Material S1). The intervention was implemented in both groups (Early-start and Delayed-start) at different time points.

For the Early-start group (nutritional counseling with a subsequent follow-up phase to evaluate carryover effects), participants allocated to this group underwent an initial in-person assessment at baseline (T0), conducted by a blinded researcher (R1). After, participants received the printed nutritional counseling material, with further explanations provided by non-blinded researchers (R2 and R3). The intervention was developed based on previous studies [5,18,19] and current dietary recommendations. In addition, during monthly remote contacts, participants were encouraged to ask questions, clarify doubts, and discuss difficulties related to dietary changes and symptom management, thereby enabling more individualized support and reinforcing the proposed guidance.

During the first three months, participants remained under follow-up and received two remote assessments through previously scheduled monthly phone calls conducted by a non-blinded researcher (R4) at T1 and T2. At the end of the intervention period (T3), participants were reassessed in person by R1 for the same outcomes evaluated at baseline. Subsequently, all active support and adherence reinforcement were discontinued, while remote assessments via phone calls at T4 and T5, conducted by R4, were maintained. At the end of the sixth month (T6), a final in-person assessment was conducted by R1 to evaluate the residual effects of the nutritional intervention.

For the Delayed-start group (delayed-start nutritional education intervention), after baseline assessment (T0) conducted by R1, participants were informed by non-blinded researchers (R2 and R3) that they would remain on a waitlist for three months and would not receive the nutritional education intervention during this period. During the waitlist period (three months), participants received two remote assessments through scheduled monthly phone calls conducted by R4 at T1 and T2. These contacts were brief and focused exclusively on assessing outcomes, without providing nutritional guidance or interactive support.

At the end of the waitlist period (T3), participants returned for a new in-person assessment conducted by R1 that evaluated the same outcomes as at baseline. Subsequently, participants received the nutritional education program counseling delivered by non-blinded researchers (R2 and R3) over the following three months, along with monthly remote assessments (T4 and T5) conducted by R4 using the same protocol as for the Early-start group. At the end of the sixth month (T6), all participants underwent a final in-person assessment conducted by R1 (Fig. 1).

### 2.4. Outcomes

The primary outcome was the number of days with at least one bowel movement during the last week (number of bowel movements/week; range 0-7). Also, stool consistency and bowel habits were assessed using the Bristol Stool Form Scale, which has been translated, adapted, and validated for the Brazilian population [14]. The scale classifies stool shape and consistency into seven categories, with types 1 and 2 indicating IC, while types 3 and 4 are considered ideal stools. Participants selected the stool type that most closely resembled their usual bowel pattern.

The main secondary outcomes were related to diet adherence. Adherence to Mediterranean diet principles was assessed using two validated instruments: an adapted Food Frequency Questionnaire (FFQ) [20] to estimate habitual dietary intake and the Mediterranean Diet Adherence Screener (MEDAS) [21]. Higher MEDAS scores indicated greater adherence to the Mediterranean dietary pattern.

While the FFQ provides a broader assessment of usual food consumption, the MEDAS is specifically designed to capture adherence to Mediterranean diet The MEDAS evaluates key components of this dietary pattern, including: abundant use of olive oil; consumption of ≥2 daily servings of vegetables; ≥2–3 daily servings of fresh fruits (including 100% natural juice); ≥3 weekly servings of legumes; ≥3 weekly servings of fish or seafood (with at least one serving of fatty fish); ≥3 weekly servings of nuts or seeds; preference for white meats over red or processed meats; and consumption of sofrito sauce at least twice per week.

During in-person assessments, data were collected on the number of bowel movements per week, stool consistency using the Bristol Stool Form Scale, and dietary intake through the FFQ and MEDAS score. During remote assessments through phone calls, the number of bowel movements per week, the Bristol Stool Form Scale, FFQ, and the MEDAS score were recorded.

### 2.5. Statistical analyses

We performed a modified intention-to-treat analysis that included all randomized participants with a baseline assessment (T0) and at least one post-baseline measurement. The primary analysis compared changes from baseline (T0) to 3 months (T3) between the Early-start and Delayed-start groups. Secondary analyses evaluated trajectories over the entire follow-up (T0 to T6). Outcomes included the number of bowel movements per week (primary outcome), the Bristol Stool Form Scale, the FFQ score, and the MEDAS score. Repeated measurements were analyzed using generalized estimating equation (GEE) models to account for within-subject correlation across visits. For the number of bowel movements per week, we fitted a binomial GEE model (7 trials per week) with a logit link, and effect estimates were shown as OR with 95% confidence intervals (95% CI). Continuous outcomes were analyzed using Gaussian GEE models with an identity link, and effect estimates were shown as β (95% CI). Models included group, time, and their interaction for between-group comparisons, and were adjusted for age and sex. Sensitivity analyses were performed to assess potential contamination in the Delayed-start group during the waitlist period.

For sample size calculation, we used data from a previous randomized controlled trial in patients with PD and IC, which reported a mean of 4.18 bowel movements per week in the intervention group and 2.81 in the control group [22]. Assuming a two-sided type I error of 0.05 and 80% power to detect a between-group difference of this magnitude, the minimum required sample size was estimated at 17 participants per group (34 total). To account for potential attrition and incomplete follow-up, we planned to recruit approximately 40 participants and allocate them 1:1 into two groups.

Baseline comparisons between groups used Fisher’s exact test for categorical variables and the Mann–Whitney test for continuous variables. Missing data were not imputed; analyses used all available observations. Statistical significance was set at two-sided p < 0.05. Statistical analyses were performed using IBM SPSS Statistics (version 23.0) and R (version 4.0.4) with the geepack and emmeans packages.

## 3. Results

We invited 43 individuals with PD and IC to participate in the study, and 38 volunteered. After inclusion, participants were allocated to the Early-start (n = 18) and Delayed-start (n = 20) groups, as shown in the study participant flow diagram (Fig. S1). For baseline data, no significant differences were observed between the Early-start and Delayed-start groups (Table 1).

**Table 1.** Baseline sociodemographic, clinical, and nutritional characteristics of participants, according to intervention group.

| **Variable<sup>a</sup>** | **Early-start (n=18)** | **Delayed-start (n=20)** | **p-value** |
| --- | --- | --- | --- |
| **Male sex, % (n)** | 50 (9) | 50 (10) | 1.000 |
| **Age<sup>a</sup>** | 64.5 (61–70) | 58 (51–64) | 0.845 |
| **MDS-UPDRS<sup>a</sup>** | 84 (69–119) | 89.5 (77–123) | 0.912 |
| **LEDD<sup>a</sup>** | 500 (387–887) | 700 (412–1132) | 0.912 |
| **Hoehn & Yahr stage $\leq 2$, % (n)** | 66.7 (12) | 75 (15) | 0.587 |
| **Marital Status – With partner, % (n)** | 61.1 (11) | 50 (10) | 0.379 |
| **Education – college level or lower, % (n)** | 61.1 (11) | 35 (7) | 0.134 |
| **Occupation - Retired, % (n)** | 72.2 (13) | 55 (11) | 0.435 |
| **Income $\leq 1$ minimum wage, % (n)** | 88.9 (16) | 60 (12) | 0.233 |
| **Disease duration $> 5$ years, % (n)** | 55.6 (10) | 50 (10) | 0.732 |
| **Practice of physical activity, % (n)** | 33.3 (6) | 60 (12) | 0.100 |
| **Hydration $\leq 1$ liter/day, % (n)** | 83.3 (15) | 55 (11) | 0.127 |
| **Gastrointestinal Symptoms** |  |  |  |
| Dysphagia, % (n) | 55.6 (10) | 50 (10) | 0.732 |
| Impaired chewing, % (n) | 44.4 (8) | 45 (9) | 0.973 |
| Dysgeusia, % (n) | 55.6 (10) | 50 (10) | 0.732 |
| Hyposmia, % (n) | 66.7 (12) | 60 (12) | 0.671 |

Abbreviations: LEDD = Levodopa Equivalent Daily Dose; MDS-UPDRS, Movement Disorder Society–Unified Parkinson’s Disease Rating Scale.
<sup>a</sup> Values in median (interquartile range).

### 3.1. Primary analysis – effects of nutritional intervention between groups after the first three months

There was no change in the number of bowel movements per week from baseline (T0) to the third month of intervention (T3) between Early-start and Delayed-start groups (group × time effect: OR 0.85, 95% CI 0.5–1.3, p = 0.490). Also, there were no changes from T0 to T3 in other outcomes (group × time effect: Bristol Scale - β 0.10, 95% CI −0.4–0.6, p = 0.732; FFQ - β 2.55, 95% CI −0.7–5.8, p = 0.124; MEDAS - β 0.71, 95% CI −0.4–1.8, p = 0.214) (Table 2). Also, there was a significant change in the number of bowel movements per week, Bristol Scale, and MEDAS from T0 to T3 in Delayed-start group (time effect: bowel movements per week - OR 2.39, 95% CI 1.7–3.2, p < 0.001; Bristol Scale - β 0.90, 95% CI 0.5–1.2, p < 0.001; MEDAS - β 0.90, 95% CI 0.1–1.6, p = 0.024) (Table 2).

**Table 2.**
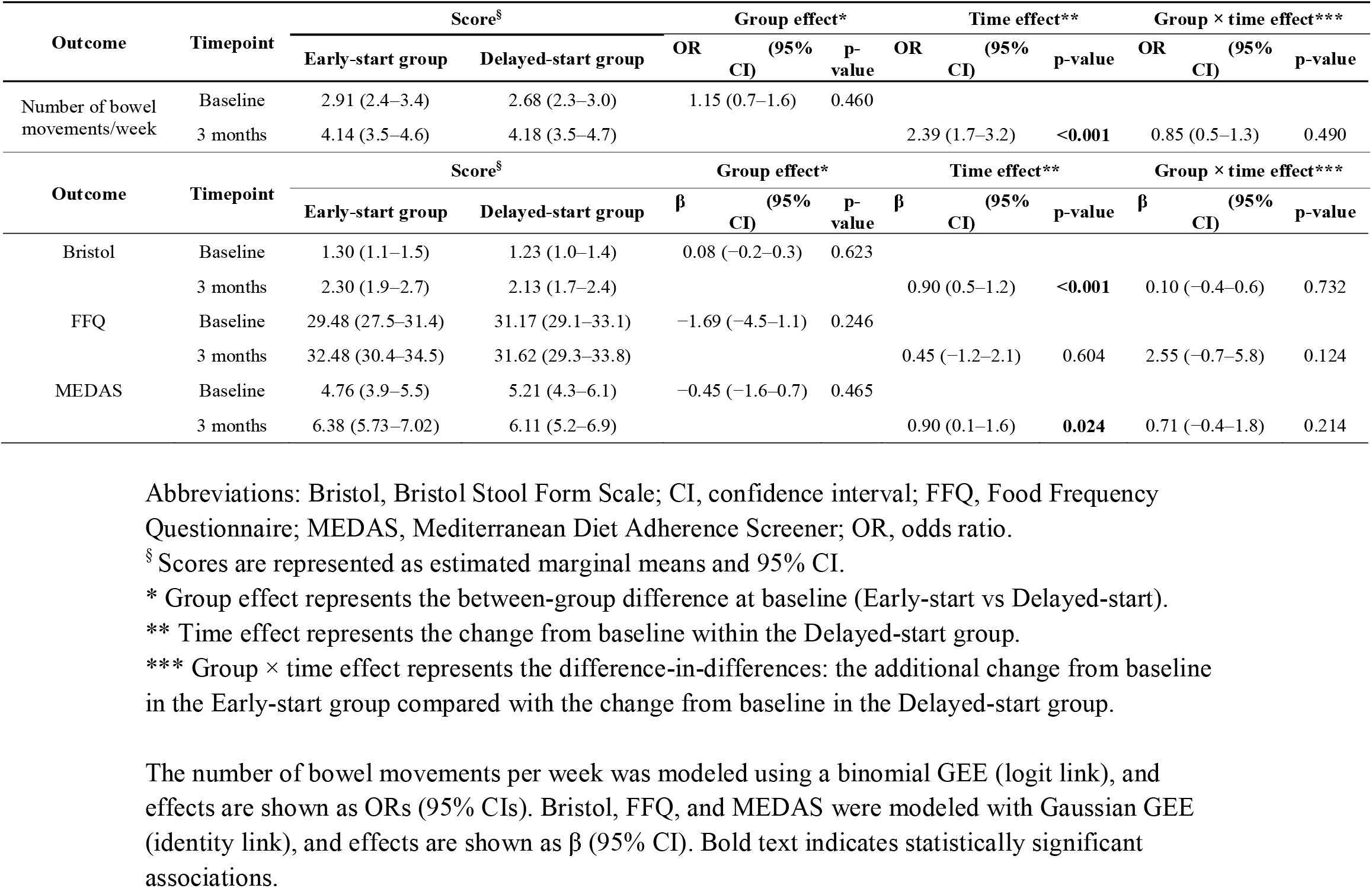
Intention-to-treat analyses of the effect of Early-start versus Delayed-start Mediterranean diet-based nutritional counseling on change from baseline clinical outcomes during the first three months of intervention in people with Parkinson’s disease and intestinal constipation.

### 3.2. Secondary analyses – longitudinal effects of nutritional intervention between groups during six months

For the Early-start group, there was a significant change in the number of bowel movements per week and normalization of stool consistency by the Bristol Scale from baseline to the sixth month (Table S1). Also, there was a progressive increase in adherence to the proposed diet, as measured by the FFQ and MEDAS, from baseline to the sixth month (Table S1). The increase in the number of bowel movements, with normalization of stool consistency, and increased adherence to the diet persisted even after the interruption of nutritional counseling in the third month of the intervention (Table S1, Fig. 2).

**Fig. 2.**
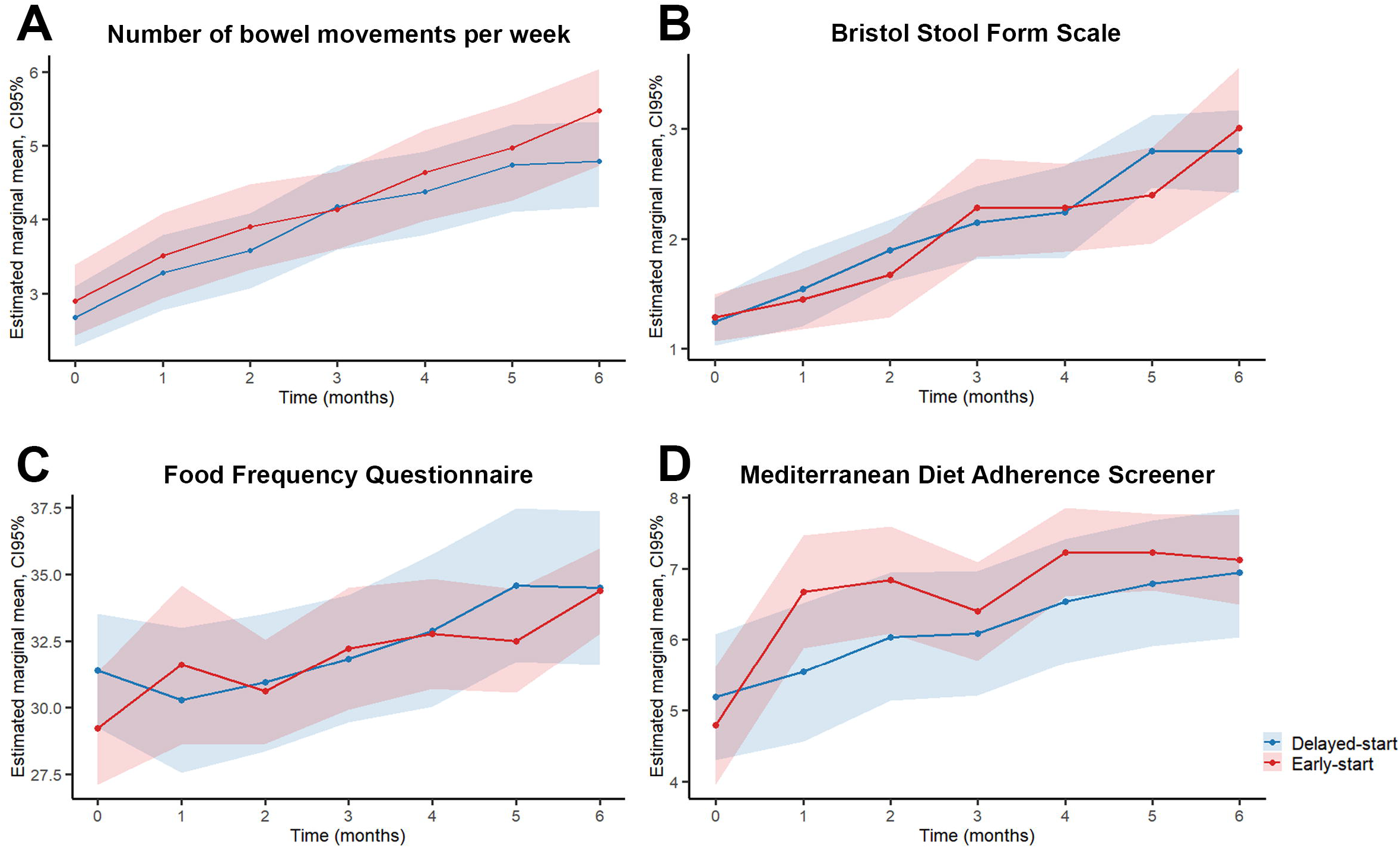
Longitudinal changes in main outcomes from baseline over six months for early-start versus delayed-start groups. A: Number of bowel movements per week (0–7 days/week). B: Bristol Stool Form Scale. C: Food Frequency Questionnaire. D: Mediterranean Diet Adherence Screener.

For the Delayed-start group, there was an unexpected increase in the number of bowel movements, with normalization of stool consistency before the start of nutritional counseling in the third month, which was also associated with adherence to the Mediterranean diet, even before the active nutritional counseling (Table S1). After the introduction of nutritional counseling, the reduction in IC symptoms continued through the sixth month, with final results similar to those of the Early-start group (Table S1, Fig. 2).

### 3.3. Sensitivity analyses

Considering the unexpected increase in the number of bowel movements, with normalization of stool consistency, and higher scores on FFQ and MEDAS in the Delayed-start group before active nutritional counseling, we performed sensitivity analyses to assess potential contamination of nutritional information for participants before active nutritional counseling. For this, we excluded participants in the Delayed-start arm who exhibited a ≥2-point increase in MEDAS from T0 to T3 (n=6), consistent with substantial dietary change during the waiting period.

After exclusion, results on the number of bowel movements per week and the Bristol score remained unchanged. However, greater adherence to the diet (FFQ, MEDAS) was observed in the Early-start group (Table S2).

## 4. Discussion

The present results showed that the proposed three-month nutritional education program for people with PD was feasible and led to adherence to the Mediterranean diet. Constipation-related outcomes improved over time during the study period, as evidenced by increased bowel movements per week and normalization of stool shape and consistency. These effects were present even three months after the conclusion of active nutritional counseling. The magnitude of these results after nutritional counseling may be clinically meaningful for people with PD. Surprisingly, some of these benefits were also observed in people with PD who did not receive the nutritional counseling (Delayed-start group), and no difference between groups was detected, which limits a causal attribution of the intervention.

Dietary interventions are an established component of IC management, particularly through increased dietary fiber intake and adequate hydration [23]. These recommendations are consistent with the Mediterranean dietary pattern, which emphasizes high intake of fruits, vegetables, legumes, and whole grains, along with healthy fats from olive oil, nuts, and seeds [24,25]. Corroborating these findings, it was described that increased intake of fiber-rich foods improved bowel movement frequency and stool consistency in individuals with chronic IC [26]. Similarly, a previous study observed improvements in IC in individuals with PD following increased adherence to a Mediterranean-style diet [5].

Dietary fiber intake is a fundamental component of IC management, and dietary patterns such as the Mediterranean diet help achieve these recommendations [23]. In this context, improving adherence to the Mediterranean diet becomes essential. Previous studies demonstrated that structured dietary counseling can effectively increase adherence to the Mediterranean diet [27,28].

Improvements in bowel habits and dietary adherence were observed over time in both groups, with no significant differences between them. This pattern suggests that changes occurred independently of group allocation. One possible explanation is the effect of study participation itself (Hawthorne effect), as regular follow-up and increased awareness may have influenced participants’ behavior. In this context, brief telephone-based contacts may have reinforced dietary modifications, as previously demonstrated in behavioral intervention studies [29], even when the focus was exclusively on assessing outcomes. In addition, participants allocated to wait-list control groups may also experience behavioral changes prior to the intervention due to expectancy effects and self-monitoring, as described previously [30]. The increase in MEDAS scores over time supports this interpretation, indicating a gradual improvement in adherence to the Mediterranean diet in both groups [20]. In addition to dietary modifications, other non-pharmacological strategies were encouraged, including increased water intake and the use of a footstool during defecation. Adequate hydration has been associated with improved stool consistency and facilitation of bowel movements [31]. Similarly, adopting a more physiological defecation posture with a footstool has been associated with reduced straining and improved bowel emptying [32].

The beneficial effects of the Mediterranean diet on IC in patients with PD may be explained by multiple physiological mechanisms. This dietary pattern is associated with modulation of the gut microbiota, promoting increased production of short-chain fatty acids, particularly butyrate, which contributes to intestinal motility and mucosal integrity [24,25]. In addition, gut health may influence the gut–brain axis, which plays a central role in the pathophysiology of PD and gastrointestinal dysfunction [6,33].

The present study has some limitations. The monocentric design may limit the generalizability of the findings, and the nature of the intervention makes double blinding difficult. The lack of strategies for monitoring spontaneous behavioral changes, particularly in the Delayed-start group, impaired recognition and avoidance of the placebo effect. The use of a delayed-start control group may introduce bias due to behavioral changes prior to the intervention. In addition, the study did not assess changes in the gut microbiota, which could provide further insight into the mechanisms underlying the observed effects. However, this is a randomized, controlled clinical trial with a single-blind design, conducted in a real-world setting of an underdeveloped region, which contributes to the development of context-specific evidence and supports the implementation of regionally tailored nutritional strategies. Also, by adapting Mediterranean diet principles into locally available foods, this low-cost educational program may be scalable to other non-Mediterranean settings and vulnerable regions. The intervention content and food-substitution list are provided in the Supplementary Material S1 to facilitate replication in other regions.

## 5. Conclusions

The results showed an improvement in bowel movement frequency and greater adherence to the Mediterranean diet during the follow-up period in individuals with PD. Although no differences were detected between the groups, our findings support Mediterranean diet–based counseling as a feasible, low-cost, and potentially scalable approach for IC management in PD when adapted to culturally relevant local foods.

## Data Availability

All data produced in the present study are available upon reasonable request to the authors

## CRediT authorship contribution statement

**Raissa Dias Fernandez:** Conceptualization, Data curation, Formal analysis, Methodology, Visualization, Writing – original draft, Writing – review & editing. **Franciele da Silva Mesquita:** Data curation, Formal analysis, Methodology. **Gabriella Cunha Ferreira:** Data curation, Formal analysis, Methodology. **Bruno Lopes Santos-Lobato:** Conceptualization, Data curation, Formal analysis, Methodology, Visualization, Supervision, Writing – original draft, Writing – review & editing.

## Consent to participate

Informed consent was obtained from all participants in the study.

## Ethics approval

The project was approved by the Research Ethics Committee of the Hospital Ophir Loyola (number 6.611.167).

## Funding disclosures for the previous 12 months

Bruno Lopes Santos-Lobato was supported by the Brazilian National Council for Scientific and Technological Development (CNPq).

## Figure Legends

Fig. S1. Study participant flow diagram. All 38 eligible participants were included in the analyses.

