## Supplementary figures and images for "Nutritional education program counseling based on the Mediterranean diet on intestinal constipation in people with Parkinson’s disease: a randomized clinical trial"

### Supplementary Figure S1

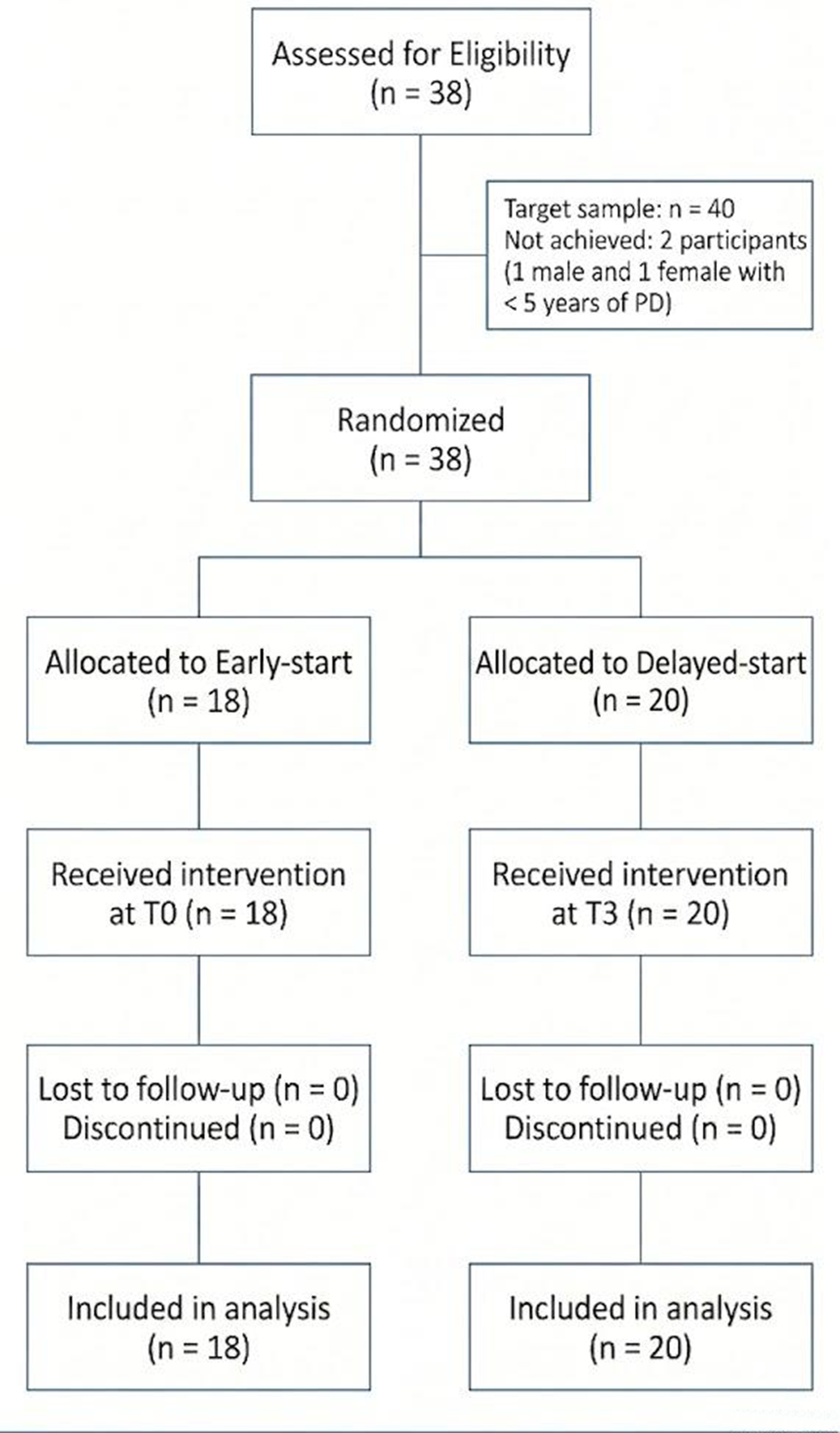
