## Supplementary Material S1 for "Nutritional education program counseling based on the Mediterranean diet on intestinal constipation in people with Parkinson’s disease: a randomized clinical trial"

[1]Laboratório de Neuropatologia Experimental, Universidade Federal do Pará, Belém, Brazil.
[2]Centro Universitário da Amazônia, Belém, Brasil.
[3]Hospital Ophir Loyola, Belém, Brazil.

**Supplementary Material S1.** Nutritional education program counseling based on the Mediterranean diet brochure for printing and distribution to participants. The brochure was provided to the Early-start group at T0 and to the Delayed-start group after the T3 visit. No nutritional guidance was provided to the Delayed-start group during the waiting period.

### GENERAL GUIDELINES FOR INTESTINAL CONSTIPATION

- Chew food thoroughly, and swallow only when it has reached a soft, paste-like consistency in the mouth.
- Whenever taking medication, use at least one full glass of water (minimum 200 mL).
- Herbal teas such as prune tea and lemon balm tea are good options for the evening.
- Psyllium is a fiber supplement that may help improve symptoms. Consume one dessert spoon per day, mixed with fruits or water. Avoid blending it, as the texture may become unpleasant. Do not exceed 7 g/day.
- The recommended daily water intake is approximately 2 liters, corresponding to 8 glasses distributed throughout the day (e.g., 4 glasses in the morning and 4 in the afternoon).

IMPORTANT: Pay attention to proper defecation posture to avoid straining and discomfort. Use a footstool to elevate the feet and try to establish a routine of sitting on the toilet at the same time each day for about 10 minutes.

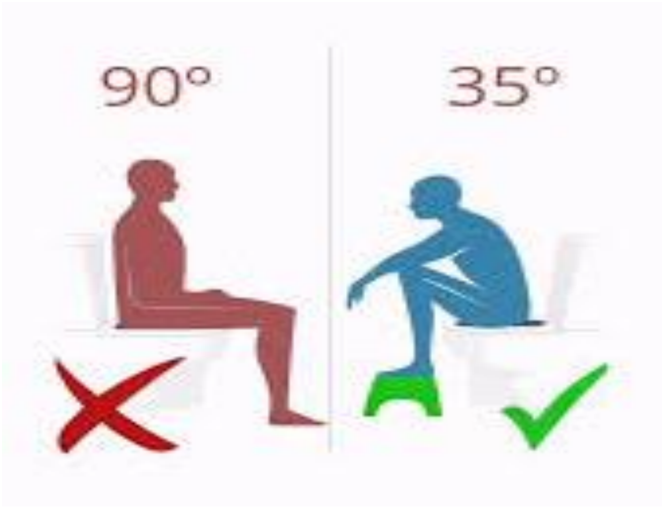

### GENERAL SUGGESTIONS FOR NORMAL DIET

- No restriction on food consistency.
- Prefer whole-grain products such as brown rice, whole-grain bread, flours, and cereals.
- Prefer raw leafy vegetables, as they are rich in fiber and help relieve constipation (e.g., lettuce, Swiss chard, kale, arugula, jambu [*an Amazonian leafy plant*], cabbage).
- When possible, add seeds to fruits or yogurt, such as chia, flaxseed, sunflower, sesame, or oats. These seeds are excellent for improving bowel function.

### SAMPLE MENU BASED ON MEDITERRANEAN DIET PRINCIPLES

**Breakfast:** Coffee with skim milk, white bread or whole-grain bread, scrambled eggs.
**Morning snack:** Fruit of choice.

**Lunch:** Raw salad with olive oil; Brown rice or white rice with vegetables (carrot, beetroot, jambu, chicory [*a bitter leafy green vegetable*], among others); Legumes (beans, preferably soaked for 8–12 hours before cooking to reduce bloating and gas issues, or chickpeas, lentils, or peas); Protein: chicken (2–3 times/week), fish (3–5 times/week), or red meat (once/week), preferably boiled, grilled, or baked.

**Dessert:** Açaí (*an Amazonian fruit*), avoid adding cassava flour, as it may worsen intestinal constipation; if possible, add oat flour, or have fruit juice.

**Afternoon snack:** Fruits.

**Dinner:** Same as lunch; avoid legumes.

**Evening snack:** Tea with whole-grain toast.

### VARIATIONS FOR MENU BASED ON MEDITERRANEAN DIET PRINCIPLES

**Breakfast:** Tapioca (*a cassava-based starch*), couscous (*a corn-based dish commonly consumed in Brazil*), bread, porridge, fruit smoothies, fruit juice.

**Morning snack:** Laxative fruits (avocado, mango, pineapple, prune, kiwi, watermelon, melon, apple with peel, pear, orange, tangerine, papaya, grapes, açaí).

**Lunch:** Salad and rice, or pasta, legumes, and protein.

**Afternoon snack:** Pupunha *(an Amazonian fruit, typically cooked)* with coffee, fruit smoothies, chopped fruits, simple cake with fruit juice (preferably using laxative fruits), whole-grain bread, boiled corn, and natural yogurt with fruit.

**Dinner:** Vegetable soup or same as lunch; avoid legumes.

**Evening snack:** Oat porridge, teas, and fruits.

### FOOD SUBSTITUTIONS BY GROUP

**Leafy greens and vegetables:** Lettuce, kale, bell pepper, chicory, arugula, cucumber, broccoli, green cabbage, red cabbage, spinach, onion.

**Vegetables:** Pumpkin, eggplant, beetroot, carrot, chayote, cauliflower, bitter melon (*jiló*), turnip, hearts of palm, okra, tomato, green beans.

**Legumes:** Black beans, carioca beans, peas, lentils, white beans, chickpeas, soybeans.

**Meat and protein sources:** Chicken, fish, eggs, or omelet, red meat (preferably boiled, baked, or grilled).

**Fats:** Avoid: cream, mayonnaise, margarine, whole milk, fried foods; Prefer: olive oil, butter.

**Breads and cereals:** Prefer whole-grain bread, whole-grain toast, oats, granola.

**Rice, pasta, and starchy foods:** White or brown rice, baked potatoes, sweet potatoes, cassava, cooked pasta, corn, whole-grain flours.

**Dairy products:** Avoid: whole milk; Prefer: semi-skimmed or skim milk, natural yogurt, mozzarella cheese, ricotta, cream cheese

**Seeds and nuts:** Cashew nuts, peanuts, almonds, Brazil nuts, walnuts, flaxseed, sesame seeds, chia.
