## Supplementary Table S1 for "Nutritional education program counseling based on the Mediterranean diet on intestinal constipation in people with Parkinson’s disease: a randomized clinical trial"

**Table S1.** Intention-to-treat analyses of the effect of Early-start versus Delayed-start Mediterranean diet-based nutritional counseling on change from baseline clinical outcomes during the intervention period (six months) in people with Parkinson’s disease and intestinal constipation.

| **Outcome** | **Timepoint** | **Score^§^** | | **Group effect*** | | | **Time effect**** | | | **Group × time effect***** | |
| --- | --- | --- | --- | --- | --- | --- | --- | --- | --- | --- | --- |
|  |  | **Early-start group** | **Delayed-start group** | **OR (95% CI)** | **p-value** | **OR (95% CI)** | | **p-value** | **OR (95% CI)** | | **p-value** |
| Number of bowel movement/week | T0 | 2.90 (2.4–3.3) | 2.68 (2.2–3.1) | 1.14 (0.7–1.6) | 0.505 |  | |  |  | |  |
|  | T1 | 3.51 (2.9–4.0) | 3.28 (2.7–3.7) |  |  | 1.42 (1.1–1.8) | | **0.008** | 1.00 (0.6–1.5) | | 0.996 |
|  | T2 | 3.91 (3.3–4.4) | 3.58 (3.0–4.0) |  |  | 1.69 (1.3–2.1) | | **< 0.001** | 1.06 (0.6–1.6) | | 0.799 |
|  | T3 | 4.13 (3.5–4.6) | 4.18 (3.5–4.7) |  |  | 2.39 (1.7–3.2) | | **< 0.001** | 0.85 (0.5–1.3) | | 0.513 |
|  | T4 | 4.64 (3.9–5.2) | 4.38 (3.7–4.9) |  |  | 2.70 (1.8–3.9) | | **< 0.001** | 1.03 (0.5–1.8) | | 0.922 |
|  | T5 | 4.97 (4.2–5.5) | 4.73 (4.1–5.2) |  |  | 3.37 (2.1–5.2) | | **< 0.001** | 1.03 (0.5–1.9) | | 0.933 |
|  | T6 | 5.48 (4.7–6.0) | 4.78 (4.1–5.3) |  |  | 3.49 (2.2–5.3) | | **< 0.001** | 1.46 (0.7–2.9) | | 0.303 |
| **Outcome** | **Timepoint** | **Score^§^** | | **Group effect*** | | **Time effect**** | | | | **Group × time effect***** | |
|  |  | **Early-start group** | **Delayed-start group** | **β (95% CI)** | **p-value** | **β (95% CI)** | | **p-value** | **β (95% CI)** | | **p-value** |
| Bristol | T0 | 1.30 (1.0–1.5) | 1.23 (0.9–1.4) | 0.07 (−0.2–0.4) | 0.682 |  | |  |  | |  |
|  | T1 | 1.47 (1.2–1.7) | 1.53 (1.1–1.8) |  |  | 0.30 (−0.0–0.6) | | 0.111 | −0.13 (−0.6–0.3) | | 0.572 |
|  | T2 | 1.69 (1.3–2.0) | 1.88 (1.5–2.1) |  |  | 0.65 (0.3–0.9) | | **<0.001** | −0.26 (−0.7–0.2) | | 0.303 |
|  | T3 | 2.30 (1.8–2.7) | 2.13 (1.8–2.4) |  |  | 0.90 (0.5–1.2) | | **<0.001** | 0.10 (−0.4–0.6) | | 0.732 |
|  | T4 | 2.30 (1.9–2.6) | 2.23 (1.8–2.6) |  |  | 1.00 (0.5–1.4) | | **<0.001** | 0.00 (−0.6–0.6) | | 1.000 |
|  | T5 | 2.41 (2.0–2.8) | 2.78 (2.4–3.1) |  |  | 1.55 (1.0–2.0) | | **<0.001** | −0.44 (−1.1–0.2) | | 0.202 |
|  | T6 | 3.02 (2.4–3.5) | 2.78 (2.4–3.1) |  |  | 1.55 (1.0–2.0) | | **<0.001** | 0.17 (−0.6–0.9) | | 0.661 |
| FFQ | T0 | 29.33 (27.3–31.3) | 31.30 (29.2–33.3) | −1.97 (−4.9–1.0) | 0.198 |  | |  |  | |  |
|  | T1 | 31.72 (28.8–34.5) | 30.20 (27.4–32.9) |  |  | −1.10 (−3.1–0.9) | | 0.297 | 3.49 (0.6–6.3) | | **0.017** |
|  | T2 | 30.72 (28.8–32.6) | 30.85 (28.3–33.3) |  |  | −0.45 (−2.4–1.5) | | 0.654 | 1.84 (−0.6–4.2) | | 0.141 |
|  | T3 | 32.33 (30.2–34.4) | 31.75 (29.5–33.9) |  |  | 0.45 (−1.2–2.1) | | 0.604 | 2.55 (−0.7–5.8) | | 0.124 |
|  | T4 | 32.89 (30.8–34.9) | 32.80 (30.0–35.5) |  |  | 1.50 (−0.9–3.9) | | 0.233 | 2.06 (−1.2–5.3) | | 0.216 |
|  | T5 | 32.61 (30.7–34.4) | 34.50 (31.8–37.1) |  |  | 3.20 (1.3–5.0) | | **<0.001** | 0.08 (−2.9–3.0) | | 0.960 |
|  | T6 | 34.50 (33.0–35.9) | 34.40 (31.7–37.0) |  |  | 3.10 (1.1–5.0) | | **0.001** | 2.07 (−1.0–5.1) | | 0.191 |
| MEDAS | T0 | 4.82 (4.0–5.6) | 5.16 (4.2–6.0) | −0.33 (−1.5–0.9) | 0.597 |  | |  |  | |  |
|  | T1 | 6.71 (5.9–7.5) | 5.51 (4.5–6.4) |  |  | 0.35 (−0.3–1.0) | | 0.325 | 1.54 (0.3–2.7) | | **0.014** |
|  | T2 | 6.88 (6.1–7.6) | 6.01 (5.1–6.9) |  |  | 0.85 (0.0–1.6) | | **0.034** | 1.21 (−0.1–2.5) | | 0.076 |
|  | T3 | 6.44 (5.7–7.0) | 6.06 (5.2–6.8) |  |  | 0.90 (0.1–1.6) | | **0.024** | 0.71 (−0.4–1.8) | | 0.214 |
|  | T4 | 7.27 (6.5–7.9) | 6.51 (5.6–7.3) |  |  | 1.35 (0.5–2.1) | | **<0.001** | 1.09 (−0.1–2.3) | | 0.084 |
|  | T5 | 7.27 (6.6–7.8) | 6.76 (5.9–7.6) |  |  | 1.60 (0.9–2.3) | | **<0.001** | 0.84 (−0.2–1.9) | | 0.139 |
|  | T6 | 7.16 (6.4–7.8) | 6.91 (6.0–7.7) |  |  | 1.75 (0.9–2.5) | | **<0.001** | 0.58 (−0.6–1.8) | | 0.350 |

Abbreviations: Bristol, Bristol Stool Form Scale; CI, confidence interval; FFQ, Food Frequency Questionnaire; MEDAS, Mediterranean Diet Adherence Screener; OR, odds ratio.

^§^ Scores are represented as estimated marginal means and 95% CI.

* Group effect represents the between-group difference at baseline (T0) (Early-start vs Delayed-start).

** Time effect represents the change from baseline within the Delayed-start group (at each follow-up time point vs T0).

*** Group × time effect represents the difference-in-differences: the additional change from baseline in the Early-start group compared with the change from baseline in the Delayed-start group (at each time point).

The number of bowel movements per week was modeled using a binomial GEE (logit link), and effects are shown as ORs (95% CIs). Bristol, FFQ, and MEDAS were modeled with Gaussian GEE (identity link), and effects are shown as β (95% CI). Bold text indicates statistically significant associations.
