## Supplementary Table S2 for "Nutritional education program counseling based on the Mediterranean diet on intestinal constipation in people with Parkinson’s disease: a randomized clinical trial"

**Table S2.** Sensitivity analyses - Intention-to-treat analyses of the effect of Early-start versus Delayed-start Mediterranean diet-based nutritional counseling on change from baseline clinical outcomes during the first three months of intervention in people with Parkinson’s disease and intestinal constipation, excluding participants from the Delayed-start group who exhibited a ≥2-point increase in MEDAS from T0 to T3 (n=6).

| **Outcome** | **Timepoint** | **Score^§^** | | **Group effect*** | | | **Time effect**** | | **Group × time effect***** | |
| --- | --- | --- | --- | --- | --- | --- | --- | --- | --- | --- |
|  |  | **Early-start group** | **Delayed-start group** | **OR (95% CI)** | **p-value** | **OR (95% CI)** | | **p-value** | **OR (95% CI)** | **p-value** |
| Number of bowel movement/week | Baseline | 2.88 (2.4–3.3) | 2.94 (2.5–3.3) | 0.96 (0.6–1.4) | 0.850 |  | |  |  |  |
|  | 3 months | 4.10 (3.5–4.6) | 4.44 (3.6–5.1) |  |  | 2.39 (1.6–3.5) | | **<0.001** | 0.85 (0.5–1.4) | 0.540 |
| **Outcome** | **Timepoint** | **Score^§^** | | **Group effect*** | | **Time effect**** | | | **Group × time effect***** | |
|  |  | **Early-start group** | **Delayed-start group** | **β (95% CI)** | **p-value** | **β (95% CI)** | | **p-value** | **β (95% CI)** | **p-value** |
| Bristol | Baseline | 1.29 (1.0–1.5) | 1.30 (1.0–1.5) | −0.02 (−0.3–0.3) | 0.928 |  | |  |  |  |
|  | 3 months | 2.29 (1.9–2.6) | 2.23 (1.8–2.5) |  |  | 0.93 (0.5–−1.3) | | **<0.001** | 0.07 (−0.5–0.6) | 0.822 |
| FFQ | Baseline | 29.49 (27.4–31.4) | 32.76 (30.9–34.5) | −3.27 (−6.0–−0.5) | **0.019** |  | |  |  |  |
|  | 3 months | 32.49 (30.4–34.5) | 31.40 (29.1–33.6) |  |  | −1.36 (−2.4–−0.3) | | **0.012** | 4.36 (1.3–7.3) | **0.004** |
| MEDAS | Baseline | 4.76 (3.9–5.5) | 5.88 (4.9–6.8) | −1.12 (−2.3–0.1) | 0.071 |  | |  |  |  |
|  | 3 months | 6.37 (5.7–7.0) | 5.74 (4.7–6.6) |  |  | −0.14 (−0.4–0.1) | | 0.403 | 1.75 (0.8–2.6) | **<0.001** |

Abbreviations: Bristol, Bristol Stool Form Scale; CI, confidence interval; FFQ, Food Frequency Questionnaire; MEDAS, Mediterranean Diet Adherence Screener; OR, odds ratio.

^§^ Scores are represented as estimated marginal means and 95% CI.

* Group effect represents the between-group difference at baseline (Early-start vs Delayed-start).

** Time effect represents the change from baseline within the Delayed-start group.

*** Group × time effect represents the difference-in-differences: the additional change from baseline in the Early-start group compared with the change from baseline in the Delayed-start group.

The number of bowel movements per week was modeled using a binomial GEE (logit link), and effects are shown as ORs (95% CIs). Bristol, FFQ, and MEDAS were modeled with Gaussian GEE (identity link), and effects are shown as β (95% CI). Bold text indicates statistically significant associations.
